# COVID-19 vaccine hesitancy in diverse groups in the UK - is the driver economic or cultural in student populations?

**DOI:** 10.1101/2021.12.14.21267773

**Authors:** Francis Drobniewski, Dian Kusuma, Agnieszka Broda, Enrique Castro-Sánchez, Raheelah Ahmad

## Abstract

Studies have identified a greater reluctance for members of the Black, Asian, and minority ethnic communities to be vaccinated against COVID-19 despite a higher probability of greater harm from COVID-19. We conducted an anonymised questionnaire-based study of students (recruiting primarily before first reports of embolic events) at two London universities to identify whether economic or educational levels were primarily responsible for this reluctance: a postgraduate core group (PGCC) n=860 and a pilot study of undergraduate medical and nursing students (n=103). Asian and Black students were 2.0 and 3.2 times (PGCC) less likely to accept the COVID vaccine than White British students. Similar findings were noted in the pilot study students. As students were studying for Masters or PhD degrees and voluntarily paying high fees, educational and economic reasons were unlikely to be the underlying cause, and wider cultural reservations were more likely. Politicians exerted a strong negative influence, suggesting that campaigns should omit politicians.

## INTRODUCTION

The behavioural responses of individuals and groups to the pandemic have been central to efforts to prevent and control viral transmission. Nonpharmaceutical interventions, including self-isolation, wearing face coverings and abiding to lock-down rules and best practice guidance, have relied heavily on the public’s acceptance and sustained behaviour change. Now, with an established technological vaccine solution, there are additional behavioural responses required. First, the vaccine is one component of protection, and other prevention behaviours still need to be practised to reduce transmission. Second, and the focus of this paper, apart from the logistics of access, there is the individual decision to be made by each of us to take up the vaccine.

Across the globe, varying levels of uptake have been reported, and some controversial methods to increase uptake have been employed from positive incentives (e.g., free sausages with vaccination in one German town, participation in lotteries in Hong Kong, Canada and the USA, direct cash in Serbia and Sweden) to sanctions for failure to be vaccinated (e.g., government of Punjab in Pakistan has employed mobile phone SIM card blocking [1]. Several countries, including the UK, are considering mandatory vaccination for social and health care workers. The different approaches can be understood in terms of the hierarchical positions on the Nuffield ladder of interventions from ‘observe and monitor’ uptake all the way up to limiting choice and the possibility of regulation, although not yet instituted anywhere [2].

While we have sizeable parts of the population across the globe unvaccinated or partially vaccinated [3], every country is trying to identify the size and key determinants of those groups who hesitate over vaccine uptake in general and COVID-19 in particular. However, before we make the leap to ‘hesitancy’ or refusal, we must be sure that barriers to access have been addressed. For example, in the US, there are reports of protracted online booking systems, complex use of language, only English documentation, and refusal at centres due to lack of personal ID [4]. Opportunity costs quickly escalate for those groups already disadvantaged – over a third of Black American households are without access to a computer or broadband, and one in five households lack access to a vehicle relying solely on public transport [4]. With the backdrop of approximately 26.1 million individuals (8.1% of the U.S. population) without any health care insurance just before the pandemic began, and 55.4% relying on employer-provided coverage [5], this means the majority are in a highly vulnerable position should they lose employment. While the COVID vaccine is free in the US, irrespective of citizenship or immigration status, if your experience of USA health care has been negative due to economic reasons then this will influence knowledge, acceptance, and trust now. Why would an illegal migrant with limited language skills believe that COVID vaccination is free if nothing else is? In contrast, National Health Systems, free at the point of access, such as in the UK, address some of these barriers and forms of exclusion, at least from a health care perspective.

Nevertheless, in the UK, as in the USA, Black, Asian, and minority ethnic (BAME) groups are financially vulnerable to working in unstable employment; many live in higher density multigenerational households and are unable to work at home, making high-risk trade-offs between isolation and work, including higher use of public transport contributing to increased risk.

Members of the BAME community have also been disproportionately affected by COVID-19, i.e., higher rates of infection, hospitalisation and death [6]. In the UK, multiple explanations have been offered for this with poverty as a root underlying cause increasing risk of transmission due to high household density in multigenerational households, zero-hours contracts prohibiting isolation and work from home [7]. Fortunately, within a year of the identification and genomic sequencing of the viral cause of COVID-19, multiple highly protective vaccines have been developed. Countries such as the UK, Israel, Bahrain, member states of the EU and the USA have rolled out highly successful vaccination programmes with significant proportions of the total adult populations covered.

In a UK survey in December 2020, vaccine hesitancy was highest among Black (odds ratio 12.96, 95% confidence interval 7.34 to 22.89), Bangladeshi, and Pakistani (both 2.31, 1.55 to 3.44) populations compared with people from a white ethnic background [8]. BAME health care workers have also shown hesitancy compared to their white coworkers [8]. Similarly, in the US, Black and Hispanic individuals were less willing than Whites to receive the COVID-19 vaccine [9,10].

Was this reluctance due to a lack of knowledge or understanding of vaccine efficacy or safety, underlying poverty preventing access and uptake or deeper cultural reasons in the BAME community perhaps rooted in historical mistrust of state bodies including the health service?

Attempts to encourage vaccine uptake will depend on an understanding of the reasons underpinning the reluctance. We attempted to better understand this through our recent analysis of the perceptions and intentions of students (including BAME students) at two London universities.

## METHODS

A cohort of 860 postgraduate students completed an anonymised questionnaire relating to COVID vaccine hesitancy (questionnaire provided in Supplementary Information 1) at two leading universities in London. The postgraduate students who were working for a higher degree, including masters or PhD students, received a specific email with an access code to the questionnaire with a follow-up reminder. They were asked about their views before and after any reports of embolic side effects emerged. In our analysis, we used February-March 2021 and April-May 2021 to identify before and after, respectively [13]. The response rate was approximately 40% (those having been sent the email and completing the questionnaire), which was expected as the timing of the questionnaire was in the run-up to exams. In addition, a pilot study of 103 undergraduate medical and nursing students was conducted by posting information on relevant physical and virtual notice boards for medical and nursing students.

The main outcome variable is vaccine acceptance. For acceptance, participants responded affirmatively (agree/strongly agree) when asked “How do you feel about the COVID-19 vaccine today?” For uptake, participants responded yes when asked “Have you had a COVID-19 vaccination?” Moreover, we asked a series of questions related to levels of confidence in the vaccine, preferred conditions (e.g., I am more likely to take the COVID-19 vaccine if:), sources of information about the vaccine, and history of influenza vaccine. We also collected socioeconomic indicators, including gender, age, ethnicity, education, and being medical or nursing students.

At the time of questionnaire completion, the cohort would not have been of an age receiving routine vaccination in the UK, but many would have been vaccinated due to professional reasons, such as being a medical student in the hospital or vaccine volunteer. We therefore included a question about COVID vaccination status.

We conducted descriptive and multivariate regression analyses. For descriptive analyses, we provided the sample characteristics and prevalence of participants who responded affirmatively (agree/strongly agree or yes). For regression analyses, we used multivariate logistic regression, controlling for socioeconomic variables. All analyses were conducted in STATA MP 15.1. We analysed the core postgraduate cohort (PGCC) as a uniform group and compared them with the pilot group of medial and nursing students where helpful.

Ethics was obtained from the Imperial College Research Ethics Committee (Ref: 21IC6546) and City University Research Ethics Committee (Ref: ETH2021-0904). Informed consent was obtained from all participants.

## RESULTS

The demographic characteristics of the full cohort of students are included in Table 1 and show that students were predominantly between 22 and 30 years of age (**Table 1**).

**Table 1.** Patient cohort sample characteristics.

|  | All postgraduates (N=860) |  | All students (N=963) |  |
| --- | --- | --- | --- | --- |
|  | n | % | n | % |
|  | [1] | [2] | [3] | [4] |
| **(a) Characteristics** |  |  |  |  |
| Gender |  |  |  |  |
| Female | 517 | 60.8 | 609 | 63.2 |
| Male | 333 | 39.2 | 342 | 35.5 |
| Other | 10 | 1.2 | 12 | 1.3 |
| Age group |  |  |  |  |
| 18-21 | 33 | 3.8 | 100 | 10.4 |
| 22-24 | 313 | 36.4 | 327 | 34.0 |
| 25-27 | 216 | 25.1 | 219 | 22.7 |
| 28-30 | 110 | 12.8 | 113 | 11.7 |
| 31-39 | 122 | 14.2 | 129 | 13.4 |
| 40+ | 66 | 7.7 | 75 | 7.8 |
| Ethnic |  |  |  |  |
| White | 540 | 62.8 | 581 | 60.3 |
| Asian | 198 | 23.0 | 232 | 24.1 |
| Black | 47 | 5.5 | 60 | 6.2 |
| Others | 75 | 8.7 | 90 | 9.4 |
| Education |  |  |  |  |
| GCSE/A level | n/a | n/a | 103 | 10.7 |
| Bachelor | 329 | 38.3 | 329 | 34.2 |
| Master/PhD | 520 | 60.5 | 520 | 54.0 |
| Other | 11 | 1.3 | 11 | 1.1 |
| Student med/nurse |  |  |  |  |
| Yes | 106 | 12.3 | 177 | 18.4 |
| No | 754 | 87.7 | 786 | 81.6 |
| Education med/nurse |  |  |  |  |
| Yes | 134 | 15.6 | 205 | 21.3 |
| No | 726 | 84.4 | 758 | 78.7 |
| **(b) COVID-19 vaccine** |  |  |  |  |
| Vaccine acceptance |  |  |  |  |
| Yes | 802 | 93.3 | 882 | 91.6 |
| No | 32 | 3.7 | 52 | 5.4 |
| Undecided | 26 | 3.0 | 29 | 3.0 |
| Got vaccine (at least one dose) |  |  |  |  |
| Yes | 252 | 29.3 | 311 | 32.3 |
| No | 608 | 70.7 | 652 | 67.7 |
| Among got vaccine, second dose |  |  |  |  |
| Yes | 124 | 49.2 | 147 | 47.3 |
| No | 128 | 50.8 | 164 | 52.7 |

Note: N or n=Observations

**Table 2** shows the level of confidence, preference, source of information, and flu vaccine history towards vaccine acceptance and uptake. For PGCC, 91% were confident that the COVID vaccines were safe (Panel a, Column 2). Belief in long-term safety was similar, as was the proportion who thought that the vaccine had been adequately tested. Overall, scientists and health care professionals had a strong positive influence on safety and efficacy perception with an equally strong negative effect when statements were made by politicians. A small percentage (7%; Panel a, Row 9, Column 2) of all respondents preferred to “have COVID-19 and develop their own immunity.”

**Table 2.** Level of confidence, preference, source of information, flu vaccine history towards vaccine acceptance and uptake.

|  | *Participants that responded affirmatively (agree/strongly agree)* |  |  |  |  |  |  |  |  |  |
| --- | --- | --- | --- | --- | --- | --- | --- | --- | --- | --- |
|  | *All respondents*<br>*N=860* |  | *Vaccine acceptance*<br>*N=802* |  | *Vaccine hesitant*<br>*N=58* |  | *Got vaccine*<br>*N=252* |  | *Not yet vaccine*<br>*N=608* |  |
|  | *n* | *%* | *n* | *%* | *n* | *%* | *n* | *%* | *n* | *%* |
|  | [1] | [2] | [3] | [4] | [5] | [6] | [7] | [8] | [9] | [10] |
| **(a) Levels of confidence in the vaccine** |  |  |  |  |  |  |  |  |  |  |
| 1. I am confident that the COVID-19 vaccine available to me is safe | 783 | 91% | 773 | 96% | 10 | 17% | 239 | 95% | 544 | 89% |
| 2. I am confident about the safety of the first batch of vaccines developed | 756 | 88% | 749 | 93% | 7 | 12% | 235 | 93% | 521 | 86% |
| 3. I am confident about the long-term safety of the vaccine offered to me | 703 | 82% | 700 | 87% | 3 | 5% | 219 | 87% | 484 | 80% |
| 4. I am concerned about the immediate/short terms side effects of the vaccine | 253 | 29% | 226 | 28% | 27 | 47% | 69 | 27% | 184 | 30% |
| 5. I think that the risk of having the vaccine is greater than the risk of COVID-19 | 92 | 11% | 70 | 9% | 22 | 38% | 26 | 10% | 66 | 11% |
| 6. I think the vaccine has been adequately tested | 717 | 83% | 707 | 88% | 10 | 17% | 220 | 87% | 497 | 82% |
| 7. I believe that the vaccine is not as good as it has been reported | 94 | 11% | 64 | 8% | 30 | 52% | 24 | 10% | 70 | 12% |
| 8. I think the vaccine would not work as well for me | 25 | 3% | 17 | 2% | 8 | 14% | 7 | 3% | 18 | 3% |
| 9. I would prefer to have COVID-19 and develop my own immunity | 61 | 7% | 40 | 5% | 21 | 36% | 18 | 7% | 43 | 7% |
| 10. I am unhappy that the second dose of vaccine is being delayed | 464 | 54% | 446 | 56% | 18 | 31% | 103 | 41% | 361 | 59% |
| 11. I do trust statements made about COVID 19 vaccine safety made by politicians | 373 | 43% | 368 | 46% | 5 | 9% | 105 | 42% | 268 | 44% |
| 12. I do trust statements made about COVID 19 vaccine safety made by scientists/doctors | 796 | 93% | 775 | 97% | 21 | 36% | 239 | 95% | 557 | 92% |
| 13. I do trust statements made about COVID 19 vaccine safety made by health care professionals (other than doctors) | 716 | 83% | 702 | 88% | 14 | 24% | 223 | 88% | 493 | 81% |
| 14. I do trust statements made about COVID 19 vaccine efficacy (how well the vaccine works) made by politicians | 394 | 46% | 389 | 49% | 5 | 9% | 111 | 44% | 283 | 47% |
| 15. I do trust statements made about COVID 19 vaccine efficacy (how well the vaccine works) made by scientists/doctors | 805 | 94% | 780 | 97% | 25 | 43% | 241 | 96% | 564 | 93% |
| **(b) I am more likely to take the Covid-19 vaccine if:** |  |  |  |  |  |  |  |  |  |  |
|  | *Participants that responded affirmatively (yes)* |  |  |  |  |  |  |  |  |  |
| 1. Available at my place of work during working hours | 261 | 88% | 231 | 89% | 30 | 77% | 151 | 89% | 110 | 86% |
| 2. Available at my GP | 265 | 87% | 240 | 90% | 25 | 68% | 148 | 86% | 117 | 89% |
| 3. I am given time off from work afterwards | 311 | 81% | 280 | 81% | 31 | 78% | 125 | 71% | 186 | 89% |
| 4. I am updated on how many staff have had the vaccine | 384 | 75% | 356 | 75% | 28 | 76% | 106 | 52% | 278 | 90% |
| 5. Colleagues from the same profession have had the vaccine. | 405 | 82% | 374 | 82% | 31 | 78% | 125 | 67% | 280 | 91% |
| 6. Colleagues from different professions have had the vaccine | 419 | 81% | 389 | 81% | 30 | 77% | 127 | 65% | 292 | 91% |
| 7. Hospital leaders/management have had the vaccine | 364 | 82% | 333 | 82% | 31 | 79% | 134 | 71% | 230 | 90% |
| 8. I have an opportunity to ask questions and think about the vaccine before making a decision | 369 | 91% | 340 | 92% | 29 | 85% | 154 | 89% | 215 | 93% |
| 9. I have enough information about the safety of the vaccine | 255 | 94% | 230 | 96% | 25 | 83% | 155 | 96% | 100 | 92% |
| 10. Initial batches of vaccine have already been used successfully | 270 | 90% | 242 | 91% | 28 | 82% | 151 | 90% | 119 | 91% |
| 11. It was Recommended by my GP | 332 | 85% | 298 | 87% | 34 | 69% | 143 | 80% | 189 | 89% |
| 12. It was recommended by a scientific expert or doctor | 249 | 91% | 226 | 95% | 23 | 64% | 152 | 94% | 97 | 87% |
| 13. It was recommended by my religious leader, e.g., priest, Imam, rabbi, etc. | 605 | 78% | 569 | 79% | 36 | 65% | 113 | 48% | 492 | 91% |
| 14. It was recommended by a celebrity (e.g., TV or film star) | 600 | 75% | 563 | 76% | 37 | 65% | 99 | 42% | 501 | 90% |
| 15. It was recommended by someone famous from my age group | 584 | 75% | 548 | 76% | 36 | 65% | 96 | 41% | 488 | 90% |
| **(c) Sources of information about the vaccine - keeping up to date** |  |  | *Participants that responded affirmatively (yes)* |  |  |  |  |  |  |  |
| 1. Official national sources | 676 | 79% | 631 | 79% | 45 | 78% | 205 | 81% | 471 | 77% |
| 2. Professional or scientific society | 654 | 76% | 608 | 76% | 46 | 79% | 203 | 81% | 451 | 74% |
| 3. Technical Sources/guidelines | 576 | 67% | 533 | 66% | 43 | 74% | 185 | 73% | 391 | 64% |
| 4. Professional network (online or in person) | 480 | 56% | 441 | 55% | 39 | 67% | 148 | 59% | 332 | 55% |
| 5. Social network (online or in person) | 375 | 44% | 342 | 43% | 33 | 57% | 102 | 40% | 273 | 45% |
| 6. Workers union | 192 | 22% | 177 | 22% | 15 | 26% | 60 | 24% | 132 | 22% |
| 7. Other Media formats | 326 | 38% | 296 | 37% | 30 | 52% | 92 | 37% | 234 | 38% |
| **(d) Out of the examples previously provided what source of information about vaccines do you trust most?** |  |  | *Each participant chose one answer* |  |  |  |  |  |  |  |
| 1. Professional or scientific society | 538 | 63% | 506 | 63% | 32 | 55% | 146 | 58% | 392 | 64% |
| 2. Official national sources | 180 | 21% | 174 | 22% | 6 | 10% | 61 | 24% | 119 | 20% |
| 3. Technical Sources/guidelines | 95 | 11% | 89 | 11% | 6 | 10% | 34 | 13% | 61 | 10% |
| 4. People, i.e., other health | 13 | 2% | 9 | 1% | 4 | 7% | 4 | 2% | 9 | 1% |
| 5. Other Media formats, i.e., Pharmaceutical | 13 | 2% | 9 | 1% | 4 | 7% | 2 | 1% | 11 | 2% |
| 6. Journalists and news | 11 | 1% | 10 | 1% | 1 | 2% | 2 | 1% | 9 | 1% |
| 7. Social media/Internet | 8 | 1% | 3 | 0% | 5 | 9% | 2 | 1% | 6 | 1% |
| 8. Organisation, i.e., Employer Workers union | 2 | 0% | 2 | 0% |  |  | 1 | 0% | 1 | 0% |
| **(e) Did you have an influenza vaccine?** |  |  | *Participants that responded affirmatively (yes)* |  |  |  |  |  |  |  |
| 1. Did you have an influenza vaccine? - Current winter (October 2020 till now) | 206 | 24% | 202 | 25% | 4 | 7% | 117 | 46% | 89 | 15% |
| 2. Did you have an influenza vaccine? - The last winter (October 2019 - March 2020) | 200 | 23% | 194 | 24% | 6 | 10% | 108 | 43% | 92 | 15% |
| 3. Did you have an influenza vaccine? - The year before (October 2018- March 2019) | 188 | 22% | 183 | 23% | 5 | 9% | 89 | 35% | 99 | 16% |
| 4. Would you like to have an influenza vaccine this year? | 270 | 44% | 261 | 46% | 9 | 18% | 54 | 43% | 216 | 44% |
| 5. Did you have an influenza vaccine? - The past 3 years | 304 | 35% | 297 | 37% | 7 | 12% | 141 | 56% | 163 | 27% |

*Note: N or n=Observations*

In general, individuals who were “vaccine hesitant” stated that they were more likely to take the COVID-19 vaccine if it were made available at the person’s place of work, if peer colleagues and hospital leaders had been vaccinated and if there was an opportunity to ask questions about the vaccine (Panel b, Column 6).

**Table 3** shows the associations between level of confidence, preference, source of information, flu vaccine history and vaccine acceptance and uptake. Having a previous influenza vaccine or current one was strongly indicative of a desire to have a COVID-19 vaccination. Those who had an influenza vaccine in any of the past three years were 6 times more likely to want the COVID-19 vaccine (Panel d, Row 5, Column 1). A positive history of prior influenza vaccination (or view on the acceptability of influenza vaccination) provides a strong indicator of the likely acceptability of COVID-19 vaccination. This group of respondents would not have been routinely offered influenza vaccine as they were too young.

**Table 3.**
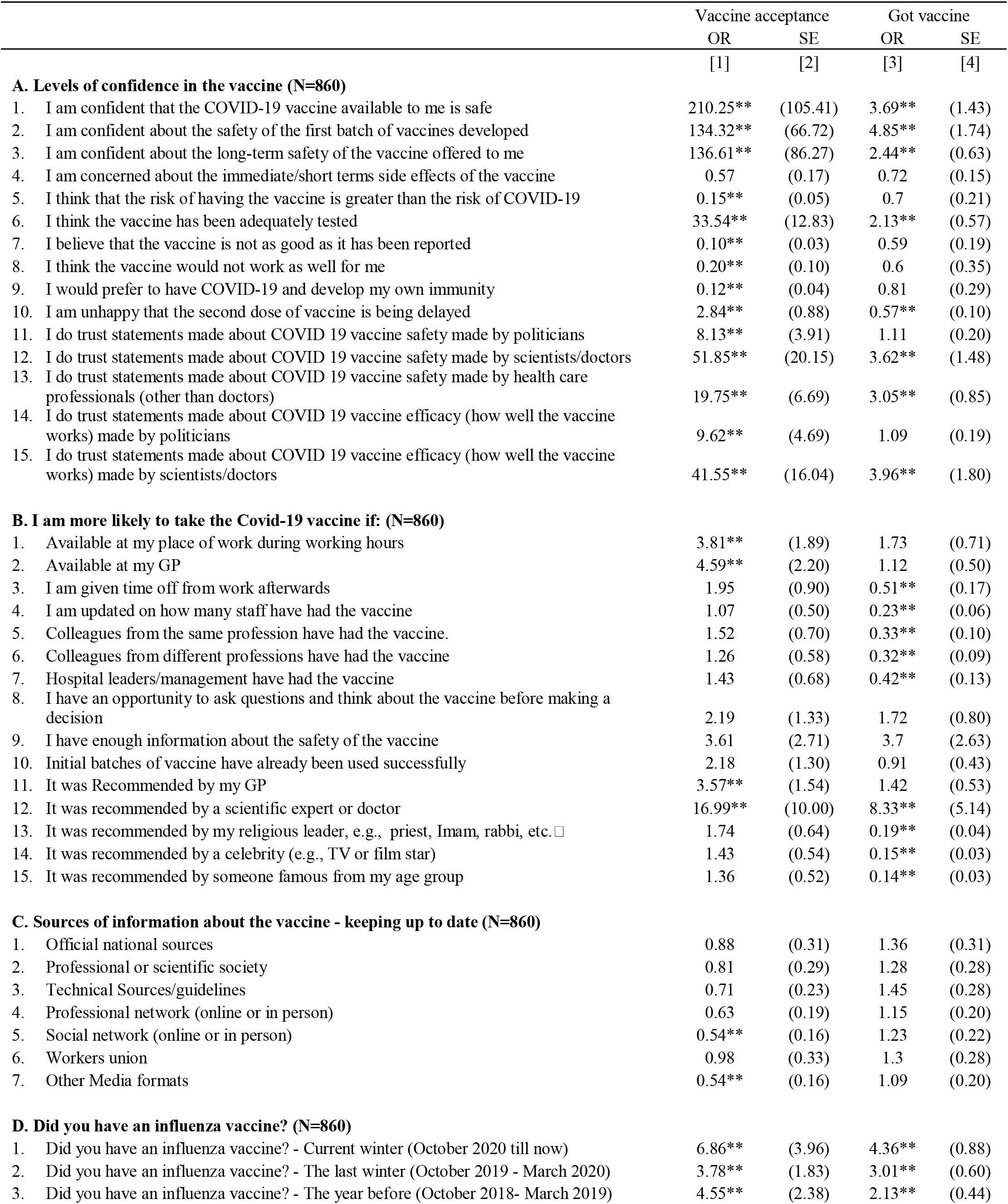
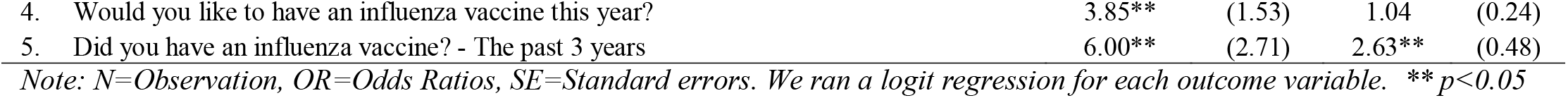
Associations between level of confidence, preference, source of information, flu vaccine history and vaccine acceptance and uptake.

|  | Vaccine acceptance |  | Got vaccine |  |
| --- | --- | --- | --- | --- |
|  | OR | SE | OR | SE |
|  | [1] | [2] | [3] | [4] |
| **A. Levels of confidence in the vaccine (N=860)** |  |  |  |  |
| 1. I am confident that the COVID-19 vaccine available to me is safe | 210.25** | (105.41) | 3.69** | (1.43) |
| 2. I am confident about the safety of the first batch of vaccines developed | 134.32** | (66.72) | 4.85** | (1.74) |
| 3. I am confident about the long-term safety of the vaccine offered to me | 136.61** | (86.27) | 2.44** | (0.63) |
| 4. I am concerned about the immediate/short terms side effects of the vaccine | 0.57 | (0.17) | 0.72 | (0.15) |
| 5. I think that the risk of having the vaccine is greater than the risk of COVID-19 | 0.15** | (0.05) | 0.7 | (0.21) |
| 6. I think the vaccine has been adequately tested | 33.54** | (12.83) | 2.13** | (0.57) |
| 7. I believe that the vaccine is not as good as it has been reported | 0.10** | (0.03) | 0.59 | (0.19) |
| 8. I think the vaccine would not work as well for me | 0.20** | (0.10) | 0.6 | (0.35) |
| 9. I would prefer to have COVID-19 and develop my own immunity | 0.12** | (0.04) | 0.81 | (0.29) |
| 10. I am unhappy that the second dose of vaccine is being delayed | 2.84** | (0.88) | 0.57** | (0.10) |
| 11. I do trust statements made about COVID 19 vaccine safety made by politicians | 8.13** | (3.91) | 1.11 | (0.20) |
| 12. I do trust statements made about COVID 19 vaccine safety made by scientists/doctors | 51.85** | (20.15) | 3.62** | (1.48) |
| 13. I do trust statements made about COVID 19 vaccine safety made by health care professionals (other than doctors) | 19.75** | (6.69) | 3.05** | (0.85) |
| 14. I do trust statements made about COVID 19 vaccine efficacy (how well the vaccine works) made by politicians | 9.62** | (4.69) | 1.09 | (0.19) |
| 15. I do trust statements made about COVID 19 vaccine efficacy (how well the vaccine works) made by scientists/doctors | 41.55** | (16.04) | 3.96** | (1.80) |
| **B. I am more likely to take the Covid-19 vaccine if: (N=860)** |  |  |  |  |
| 1. Available at my place of work during working hours | 3.81** | (1.89) | 1.73 | (0.71) |
| 2. Available at my GP | 4.59** | (2.20) | 1.12 | (0.50) |
| 3. I am given time off from work afterwards | 1.95 | (0.90) | 0.51** | (0.17) |
| 4. I am updated on how many staff have had the vaccine | 1.07 | (0.50) | 0.23** | (0.06) |
| 5. Colleagues from the same profession have had the vaccine. | 1.52 | (0.70) | 0.33** | (0.10) |
| 6. Colleagues from different professions have had the vaccine | 1.26 | (0.58) | 0.32** | (0.09) |
| 7. Hospital leaders/management have had the vaccine | 1.43 | (0.68) | 0.42** | (0.13) |
| 8. I have an opportunity to ask questions and think about the vaccine before making a decision | 2.19 | (1.33) | 1.72 | (0.80) |
| 9. I have enough information about the safety of the vaccine | 3.61 | (2.71) | 3.7 | (2.63) |
| 10. Initial batches of vaccine have already been used successfully | 2.18 | (1.30) | 0.91 | (0.43) |
| 11. It was Recommended by my GP | 3.57** | (1.54) | 1.42 | (0.53) |
| 12. It was recommended by a scientific expert or doctor | 16.99** | (10.00) | 8.33** | (5.14) |
| 13. It was recommended by my religious leader, e.g., priest, Imam, rabbi, etc. □ | 1.74 | (0.64) | 0.19** | (0.04) |
| 14. It was recommended by a celebrity (e.g., TV or film star) | 1.43 | (0.54) | 0.15** | (0.03) |
| 15. It was recommended by someone famous from my age group | 1.36 | (0.52) | 0.14** | (0.03) |
| **C. Sources of information about the vaccine - keeping up to date (N=860)** |  |  |  |  |
| 1. Official national sources | 0.88 | (0.31) | 1.36 | (0.31) |
| 2. Professional or scientific society | 0.81 | (0.29) | 1.28 | (0.28) |
| 3. Technical Sources/guidelines | 0.71 | (0.23) | 1.45 | (0.28) |
| 4. Professional network (online or in person) | 0.63 | (0.19) | 1.15 | (0.20) |
| 5. Social network (online or in person) | 0.54** | (0.16) | 1.23 | (0.22) |
| 6. Workers union | 0.98 | (0.33) | 1.3 | (0.28) |
| 7. Other Media formats | 0.54** | (0.16) | 1.09 | (0.20) |
| **D. Did you have an influenza vaccine? (N=860)** |  |  |  |  |
| 1. Did you have an influenza vaccine? - Current winter (October 2020 till now) | 6.86** | (3.96) | 4.36** | (0.88) |
| 2. Did you have an influenza vaccine? - The last winter (October 2019 - March 2020) | 3.78** | (1.83) | 3.01** | (0.60) |
| 3. Did you have an influenza vaccine? - The year before (October 2018- March 2019) | 4.55** | (2.38) | 2.13** | (0.44) |
| 4. Would you like to have an influenza vaccine this year? | 3.85** | (1.53) | 1.04 | (0.24) |
| 5. Did you have an influenza vaccine? - The past 3 years | 6.00** | (2.71) | 2.63** | (0.48) |

*Note: N=Observation, OR=Odds Ratios, SE=Standard errors. We ran a logit regression for each outcome variable. \*\* $p < 0.05$*

The majority, as expected, learned about COVID vaccination mainly from professional or scientific sources, but interestingly, with limited input from other media, including social media, despite the age profile of the group (**Table 2, Panel d, Column 2**).

Considering the correlates of vaccine acceptance (**Table 4**), older age was positively associated with vaccine acceptance both before and after revelations of embolic side effects of the AstraZeneca vaccine (which subsequently led to non-AstraZeneca vaccine being chosen for younger age groups in the UK).

**Table 4.** Sociodemographic correlates of vaccine acceptance (including before/after embolism issues) and uptake.

|  | Outcome: Vaccine acceptance |  |  |  |  |  | Outcome: Got Vaccine |  |
| --- | --- | --- | --- | --- | --- | --- | --- | --- |
|  | All study period |  | Feb-Mar 2021 |  | Apr-May 2021 |  | OR | SE |
|  | OR | SE | OR | SE | OR | SE |  |  |
|  | [1] |  | [2] |  | [3] |  | [4] |  |
| Gender |  |  |  |  |  |  |  |  |
| Female | Ref. |  |  |  |  |  |  |  |
| Male | 1.41 | (0.45) | 1.36 | (0.45) | - |  | 0.55*** | (0.11) |
| Other | 0.10*** | (0.07) | 0.13** | (0.11) | - |  | 0.43 | (0.41) |
| Age group |  |  |  |  |  |  |  |  |
| 18-21 | Ref. |  |  |  |  |  |  |  |
| 22-24 | 2.38 | (1.39) | 2.34 | (1.58) | 1.82 | (2.80) | 0.83 | (0.39) |
| 25-27 | 2.34 | (1.44) | 2.14 | (1.50) | 5.65 | (10.47) | 1.37 | (0.65) |
| 28-30 | 2.23 | (1.50) | 1.86 | (1.38) | - |  | 1.88 | (0.95) |
| 31-39 | 1.95 | (1.20) | 1.71 | (1.21) | 3.98 | (7.14) | 4.08*** | (1.96) |
| 40+ | 6.15** | (4.92) | 2.61 | (2.42) | 31.93* | (61.88) | 17.74*** | (9.87) |
| Ethnicity |  |  |  |  |  |  |  |  |
| White | Ref. |  |  |  |  |  |  |  |
| Asian | 0.50** | (0.17) | 0.48** | (0.17) | 0.85 | (1.01) | 0.91 | (0.20) |
| Black | 0.31** | (0.16) | 0.32* | (0.20) | 0.19 | (0.24) | 1.42 | (0.58) |
| Other | 0.52 | (0.24) | 0.49 | (0.24) | - |  | 0.68 | (0.22) |
| Education |  |  |  |  |  |  |  |  |
| Bachelor | Ref. |  |  |  |  |  |  |  |
| Master/PhD | 0.55* | (0.19) | 0.67 | (0.26) | 0.29 | (0.28) | 0.46*** | (0.09) |
| Other | 0.21** | (0.16) | 0.18 | (0.20) | 0.08 | (0.13) | 0.43 | (0.35) |
| Medical/nursing student |  |  |  |  |  |  |  |  |
| No | Ref. |  |  |  |  |  |  |  |
| Yes | 0.52* | (0.20) | 0.55 | (0.26) | 0.72 | (0.64) | 3.06*** | (0.75) |
| Constant | 14.10*** | (8.57) | 13.07*** | (9.32) | 11.11 | (17.40) | 0.38** | (0.17) |
| N | 860 |  | 709 |  | 111 |  | 860 |  |

Note: N=Observation, OR=Odds Ratios, SE=Standard errors. \*\*\* $p < 0.01$ , \*\* $p < 0.05$ , \* $p < 0.1$

If one considers the entire cohort (i.e., the PG core plus the undergraduate medical and nursing students from the pilot study), similar trends were seen. Asian and Black students were 1.8x and 5x less likely to accept COVID vaccination compared to white British students in the total cohort and were 2.0 and 3.2x less likely in the PG core cohort. Curiously, medical and nursing students were 1.92 and 3.06 times less willing to be vaccinated than other students. This willingness to be vaccinated needs to be viewed in the context of the findings that the medical and nursing students were 2.8 times more likely to have received the vaccine at the time of the survey. For the medical/nurse student group, it would appear that although there was a collective reluctance to be vaccinated, there was pragmatic acceptance.

## DISCUSSION

The key observation was that Asian and Black students were 2.0x and 3.2x LESS likely to accept the COVID vaccine compared to White British students. The same ethnic group findings were noted in those recruited before reports of embolisms (up to 31 March 2021) and those, albeit a smaller sample, completing the questionnaire afterwards (up until 30 May).

We also explored the main sources of information on vaccine safety and efficacy in the study population, as this would be the key to influencing their views and opinions later on. It was clear that scientists/doctors had a strong positive influence on vaccine uptake, while politicians exerted a strong negative influence across *all groups*. Our findings strongly suggest that campaigns to increase vaccine confidence in BAME individuals in particular should therefore omit politicians.

In relation to the influenza vaccine, those who have had influenza vaccine in any of the past three years were 6.5 times more likely to want the COVID vaccine compared to those who have not had an influenza vaccine. Influenza vaccination is a useful marker for COVID-19 vaccination, i.e., generally supportive attitude to vaccination in general.

In this population group, knowledge of science, health and vaccines can be assumed to be high given that all participants have a bachelor’s degree and are studying for a master’s or PhD degree in health or medical sciences. We can rule out lack of knowledge/understanding as a major factor in vaccine hesitancy.

Although no direct questions were made regarding wealth, these postgraduate students voluntarily attended and paid for high-cost courses (range £15,000 to over £30,000). Within this group, we can conclude that the reasons some BAME groups are hesitant to be vaccinated cannot be due to lack of knowledge or because of poverty. Other factors, including deep held cultural beliefs or social norms as well as prior experiences with health care or health care services, may be crucial determinants.

Our study conclusions are supported by those of Sturgis et al. (2021), who used pre-COVID cross-sectional pandemic data from the Wellcome Global Monitor and showed that in countries where trust in science is high, people are also more confident about vaccination, accounting for their own level of trust in science. Countries where the consensus is that science and scientists can be trusted are high showed a positive association between that trust in science and vaccination confidence [12].

The specific findings in the pilot study demonstrated similar findings, which would need verification through a larger study. However, this group did suggest that even trainee doctors and nurses would not automatically support COVID vaccination despite arguably being closer to the effects of the virus (patient deaths, largely greater work exposure). Worryingly, with 1.3 million NHS staff, this group may have a wider negative influence against vaccination amongst the general population as well.

If compulsory vaccination of NHS and social care staff is mandated, as currently proposed in the UK, there is a risk of a negative impact on NHS staff recruitment and retention. Although the percentage staff lost would probably be small, this would be numerically significant in a workforce, and the size of the NHS would add to an existing shortfall of frontline clinical staff. If we accept that the policy is correct, then we must develop practical strategies that promote clinical staff retention against the policy background of compulsory vaccination. Table 5 gives a summary of factors that are likely to have a positive effect on COVID 19 vaccination, but which would need to be verified in a larger cohort of NHS staff.

**Table 5.** Factors that should be incorporated in all health care and social care worker COVID-19 vaccination campaigns.

|  |
| --- |
| ☐ Recommendations and promotion made by scientists, doctors and health care workers<br>☐ No statements made by politicians<br>☐ Recommendations by GPs and religious leaders helpful<br>☐ Vaccine availability at place of work during normal working hours, i.e., minimal friction to maximise vaccine uptake<br>☐ Opportunity to ask questions regarding the vaccine<br>☐ Vaccine campaigns which build on influenza vaccine campaigns<br>☐ Consider positive incentives/rewards |

We accept that as the impact of COVID-19 may not be homogeneous across diverse ethnic groups, no single communication and engagement intervention may be effective in influencing behaviours in all communities. However, we identified positive (e.g., scientist) and negative influencers (e.g., politicians) for all groups. We believe this study will help to better tailor campaigns to increase vaccine uptake where needed and further inform existing initiatives aimed at all adults [6]. Close monitoring of uptake and learning for future campaigns will be essential to ensure that all ethnic groups are able and willing to be vaccinated. When low- and middle-income countries (LMICs) are unable to source sufficient vaccine doses despite great need every behavioural strategy needs to be deployed to maximise uptake in countries which can afford more doses than their entire population. There may also be more similarities than differences between high-income and low-income settings in terms of behaviours and trusted sources; a recent study shows that health workers are the most trusted sources of guidance about COVID-19 vaccines in LMICs [13].

Similarly, vaccine hesitancy during medical and nursing training should be addressed and arguably even beforehand during high school. As the UK faces complex decisions around release from lockdown and increasing case numbers, we need to consider vaccination of teenagers (who carry and transmit but are largely immune to the lethal effects of the disease) and so family, student and teenager understanding and acceptance of vaccination both for individual health and for wider public health.

A potential weakness of our study was that we did not capture socioeconomic status, which might be a confounder within the medical and nursing groups. Nonetheless, these findings provide useful insight into disparities in uptake in health care workers and provide opportunities for earlier interventions. For example, there may be implications for how we teach microbiology/infectious diseases literacy on our medical ad nursing and other health-related courses. Understanding technology/vaccine development and safety may also be needed. There may be major implications as these students qualify and progress as health care professionals for vaccine uptake amongst the professional groups as well as the messages they relay to patients and public at large.

## Data Availability

All data produced in the present study are available upon reasonable request to the authors.

https://imperial.eu.qualtrics.com/jfe/form/SV_0xilN8FYXeabREW

## Author contributions

FD and RA conceived the study and with ECS and AB developed, tested and implemented the survey. AB also provided project management support. DK carried out the analysis with input from FD, RA and ECS. FD, RA, DK developed the manuscript. All edited and finalized the manuscript.

## Additional Information

There are no conflicts of interest. All methods were carried out in accordance with relevant guidelines and regulations for primary survey research (see ethics above).

The study was partially funded by Horizon Europe grant No. 101046016 — EuCARE: European Cohorts of Patients and Schools to Advance Response to Epidemics. FD is supported by the NIHR Imperial Biomedical Research Centre. RA, as Knowledge Mobilisation Lead, acknowledges the National Institute for Health Research Health Protection Research Unit (NIHR HPRU) in Healthcare Associated Infections and Antimicrobial Resistance at Imperial College London in partnership with the UK Health Security Agency (previously PHE), and in collaboration with, Imperial Healthcare Partners, University of Cambridge and University of Warwick. The views expressed in this article are those of the author(s) and not necessarily those of the NIHR or the NHS.

## Supplementary File

<u>COVID Vaccine perceptions survey - Drobnieweski et al</u>

## Notes

### Competing Interest Statement

The authors have declared no competing interest.

### Summary of Updates

Apologies - spotted an in correct sentence in the recently updated acknowledgements section under additional information. No other changes to the content.

